# SARS-CoV-2 Seroprevalence Among All Workers in a Teaching Hospital in Spain: Unmasking The Risk

**DOI:** 10.1101/2020.05.29.20116731

**Authors:** M Isabel Galán, María Velasco, M Luisa Casas, M José Goyanes, Gil Rodríguez-Caravaca, Juan Emilio Losa, Carmen Noguera, Virgilio Castilla, Working Group Alcorcon COVID-19 investigators

**Affiliations:** Ocupation Health Unit; Infectious Diseases. Internal Medicine Unit; Research Unit; Laboratory Unit; Microbiology Unit; Preventive Medicine Unit; Nurse Subdirector; Medical Director; Working Group Alcorcon COVID-19 investigators; Hospital Universitario Fundación Alcorcón, Alcorcón, Madrid Calle Budapest nº1, 28922 Alcorcón, Madrid. Spain

**Author notes:** Algora Weber, Alejandro; Alonso Punter, Juan Carlos; Alonso Salazar, María Teresa; Bonilla Zafra, Gregorio; Bueno Campaña, M^a^ Mercedes; Carrión Pulido, Camilo; Diaz Cuasante, Ana Isabel; Fabero Jiménez, Aurora; Fariña García, Rosa María; González Anglada, María Isabel; Guijarro Herraiz, Carlos; Izquierdo Patron, Mª Mercedes; Lorenzo Martínez, Susana; Mosquera González, Margarita; Pérez Encinas, Montserrat; Pérez Fernandez, Elia; Pérez Vega, Francisco José; Renilla Sánchez, Maria Esther. These authors contributed equal to the manuscript. **Corresponding author:** María Velasco, MD, DTMH, PhD Infectious Diseases, Internal Medicine. Research Unit.; Hospital Universitario Fundación Alcorcón C/Budapest nº1, 28922 Alcorcón. Madrid. Spain 830 430.

## Abstract

**Background:** Health-care workers (HCW) are at increased risk for SARS-CoV-2 infection, but few studies have evaluated prevalence of antibodies against SARS-CoV-2 among them.

**Objective:** To determine the seroprevalence against SARS-CoV-2 in all HCW.

**Methods:** Cross-sectional study (April 14^th^- 27^th^, 2020) of all HCW at Hospital Universitario Fundación Alcorcón, a second level teaching hospital in Madrid, Spain. SARS-CoV-2 IgG was measured by ELISA. HCW were classified by professional category, working area, and risk for SARS-CoV-2 exposure.

**Results:** Among 2919 HCW, 2590 (90.5%) were evaluated. Mean age was 43.8 years (SD 11.1) and 73.9% were females. Globally, 818 (31.6%) workers were IgG positive, with no differences for age, sex or previous diseases. Among them, 48.5% did not report previous symptoms. Seropositivity was more frequent in high (33.1%) and medium (33.8%) than in low-risk areas (25.8%, p = 0.007), but no difference was found for hospitalization areas attending COVID-19 and non-COVID-19 patients (35.5 vs 38.3% p = NS). HCW with a previous SARS-CoV2 PCR positive test were IgG seropositive in 90.8%. By multivariate logistic regression analysis, seropositivity was associated with being physicians (OR 2.37, CI95% 1.61–3.49), nurses (OR 1.67, CI95% 1.14–2.46), or nurse- assistants (OR 1.84, CI95% 1.24–2.73), HCW working at COVID-19 hospitalization areas (OR 1.71, CI95% 1.22–2.40), non-COVID-19 hospitalization areas (OR 1.88, CI95% 1.30–2.73), and at the Emergency Room (OR 1.51, CI95% 1.01–2.27)

**Conclusions:** Seroprevalence uncovered a high rate of infection previously unnoticed among HCW. Patients not suspected of having COVID-19 as well as asymptomatic HCW may be a relevant source for nosocomial SARS-CoV-2 transmission.

## INTRODUCTION

COVID-19 is a disease caused by a new human coronavirus (SARS-CoV-2) that emerged in Wuhan, China, in late 2019^1^. In Spain, the first case of SARS-CoV-2 was identified on January 31^st^ imported from Germany. Since that time, a sharp increase in the number of cases has pushed the capacity of healthcare system in Madrid beyond the limit^2^. More than 60.000 patients were attended in Madrid’s hospitals during March and April 2020^3^. As the pandemic accelerated, access to personal protective equipment (PPE) for health workers was a key concern, moreover because of PPE shortages^4^.

Health-care workers (HCW) are at increased risk for infection, and specific requirements for their protection are advisable to ensure the functioning of the healthcare system^5^. Indeed, in Spain, more than 26.000 health care workers have been infected and at least 41 had died^4^. Alongside concerns for the healthcare workers personal safety, anxiety about transmitting the infection to their relatives and patients adds another stress to HCW.

At this time, it is known that SARS-CoV-2 human to human transmission occurs during the presymptomatic stage through droplets or direct contact^6–8^. The possibility of presymptomatic transmission increases the challenges of containment measures^9–11^. Moreover, according to two studies, presumed hospital-related transmission of SARS-CoV-2 was suspected in 41% and 35% of patients^10,12,13^. Nosocomial transmission may originate from patients (where protective measures are usually strict), but also equipment (PPE) for health workers was a key concern, moreover because of PPE the presymptomatic stage from asymptomatic HCW (where protective measures may be more relaxed or simply non-existing).

Little is known about hospital HCW seroprevalence for SARS-CoV-2. Rates from other coronavirus epidemic such as MERS and SARS range between 2.3% and 20% of subclinical infection^14–16^. Nosocomial transmission has been recognized as an important amplifier in epidemics of both SARS and Middle East respiratory syndrome^17–19^.

Serological surveillance of exposed individuals allows to estimate the individual risk. This approach is essential since the safety of health-care workers must be ensured. Screening all health-care workers for SARS-CoV-2 in the hospital would be helpful to maintain the welfare of the staff and to enable identification of infected health-care workers.

Our objective was to evaluate the prevalence of immunoglobulin G (IgG) against SARS-CoV-2 among all the employees of a second level teaching hospital in the south of Madrid.

## MATERIALS AND METHODS

Design. Cross sectional study of all hospital workers, direct hospital employees (clinical and not clinical), as well as workers for external contractors developing their regular tasks inside the hospital.

Setting. HUFA is a 402 beds general hospital, including 93 internal medicine and 16 critical care beds. As the only public hospital in Alcorcon, it covers a population of 170.000 inhabitants. On March 3^th^ we received the first COVID-19 patient. Total number of COVID-19 patients attended by April 14th was 1,638 patients, among them 236 died. A quick structural and functional reorganization was required, reaching a peak occupancy of 370% in internal medicine hospitalization, 293% in ICU and 320% in ER. The number of PPE providers increased 10 fold and associated expenditures 300-fold. In spite of this, occasional shortage of appropriate PPE occurred several times.

For the purpose of the analysis, workers were classified into categories according to the estimated risks for nosocomial exposure to SARS-CoV2. Very high included professionals with direct contact with COVID-19 patients: critical care & anesthesiology, ER (ER) and COVID-hospitalization ward. Medium risk was attributed to professionals attending patients not suspected having COVID-19 both in hospital wards and outpatient clinics, and central units (pharmacy, laboratory, radiology, pathology). Finally, low risk was attributed to administrative and management units and external workers (cooks, food service, cleaners, ambulance drivers, store sellers, and watchmen)^20^ PPE were distributed according to the estimated risk in different areas, with a priority for critical care unit and ER in case of shortage. A total of 1,561 workers most likely to attend COVID-19 patients received intensive training for the use of PPE at the hospital’s Center for Medical Simulation IDEhA.

### Selection and participation of individuals

All HCW were invited to attend to an interview conducted by the staff of the Occupational Health Unit (OHU) and additional clinical assistants and blood sample extraction for serologic studies April 14^th^ to April 27^th^

All along the study period, professionals with symptoms suggestive of COVID-19 were encouraged to attended to the OHU where a nasopharyngeal swab was obtained for SARS-CoV-2 PCR exam. Patients with a positive test were sent home for quarantine or to the ER for further clinical evaluation.

Results of IgG status were informed personally to all HCW by OHU staff one to two weeks after blood extraction. HCW with a positive IgG and COVID-19 compatible symptoms in the previous 14 days were identified and tested by PCR.

### Variables

We collected information about age, gender, professional category, area of work, relocation during COVID-19 care (if appropriate) previous health conditions (Supplement 1), self-reported potential SARS-CoV-2 exposure and type of exposure, (occupational with PPE, occupational without PPE or non-occupational), last date of exposure; presence and date of COVID-19 symptoms, PCR test. Severity of disease: out-patient evaluation, ER consultation, hospital admission and clinical outcome.

### Laboratory procedures

We measured serum IgG antibody by an enzyme-linked immunosorbent assay (ELISA) IgG2 using a SARS-CoV-2 S spike and Nucleocapsid recombinant antigens (Diapro (Palex), Italy), to screen for the presence of human anti-SARS-CoV-2 IgG. Thisassay (ELISA) IgG2 using a assay (CE approved) was used according to the manufacturer’s protocol. Reported sensitivity of the assay by the manufacturer was 98% (Supplement 2)

For molecular diagnosis of SARS-CoV-2 infection of nasopharyngeal swabs were processed by automatized extraction using the MagNa Pure Lc instrument (Roche Applied Science, Mannheim, Germany) and real time reverse transcription polymerase chain reaction using the SARS-Cov-2 nucleic acid detection Viasure kit (CerTest Biotec S.L.), following the manufacturer’s instructions (Supplement 1)

### IgG results interpretation

As per hospital protocol, all samples corresponding to whole HCW were analyzed. HCW with positive IgG and presence of symptoms older than 14 days were assumed to be infected but no longer contagious. Those with positive IgG and symptoms in the past 14 days were considered as active infected and potential contagious and underwent PCR examination. If PCR results were positive, they were discharged. HCW with IgG negative were considered susceptible to SARS-CoV-2 infection^21^. Asymptomatic workers were not routinely tested with PCR, but such test was performed for persons with self-reported symptoms, and the report was voluntary.

### Statistical analysis

Data are reported as mean (± SD), median (IQR) or percentage as appropriate. Categorical variables were compared using Pearson’s X2 test or Fisher exact test. Continuous variables were analyzed using the Student t-test.

A univariate analysis was carried out to find independently associated risk factors for positive IgG. A multivariate logistic regression model evaluated the association between risk factors and positive IgG was assessed by reference to odds ratio (OR). Statistical analysis was performed, with hypothesis testing based on a two-tailed test of significance and we considered statistical significance P< 0.05 with the Statistical Package for Social Sciences (SPSSPC v 20 Illinois USA)

### Study approval / Ethics

All participants enrolled into the study voluntarily, and written informed consent was required to use the data for analysis. Participation in the study or results were not reported to the employer. The study protocol was approved by the HUFA independent ethics research committee (reference number 20/69). We stated that results not would be used to generate an immunological passport in the hospital^22^.

## RESULTS

All 2,919 HCW HUFA were invited to participate in the study between April 14–27, 2020. Among them 278 (9.5%) workers did not come to be tested because sick leave, work at home, or declined the invitation (figure 1). In addition, 51 HCW (1.8%) refused consent to use their data for investigational purposes and were removed from the analysis. Thus, data of a total of 2,590 (98%) HCW were analyzed. They were 1,915 females, (73.9%) and mean age was 43.8 (SD 11.1) years. Previous relevant clinical condition was present in 998 HCW (38.5%), distributed as follows: tobacco use 21%, chronic lung disease or asthma 8%, obesity 6.0%, high blood pressure 6.9%, diabetes mellitus 2.1% and other cardiovascular diseases 2.0% (Table 1).

**Figure 1.**
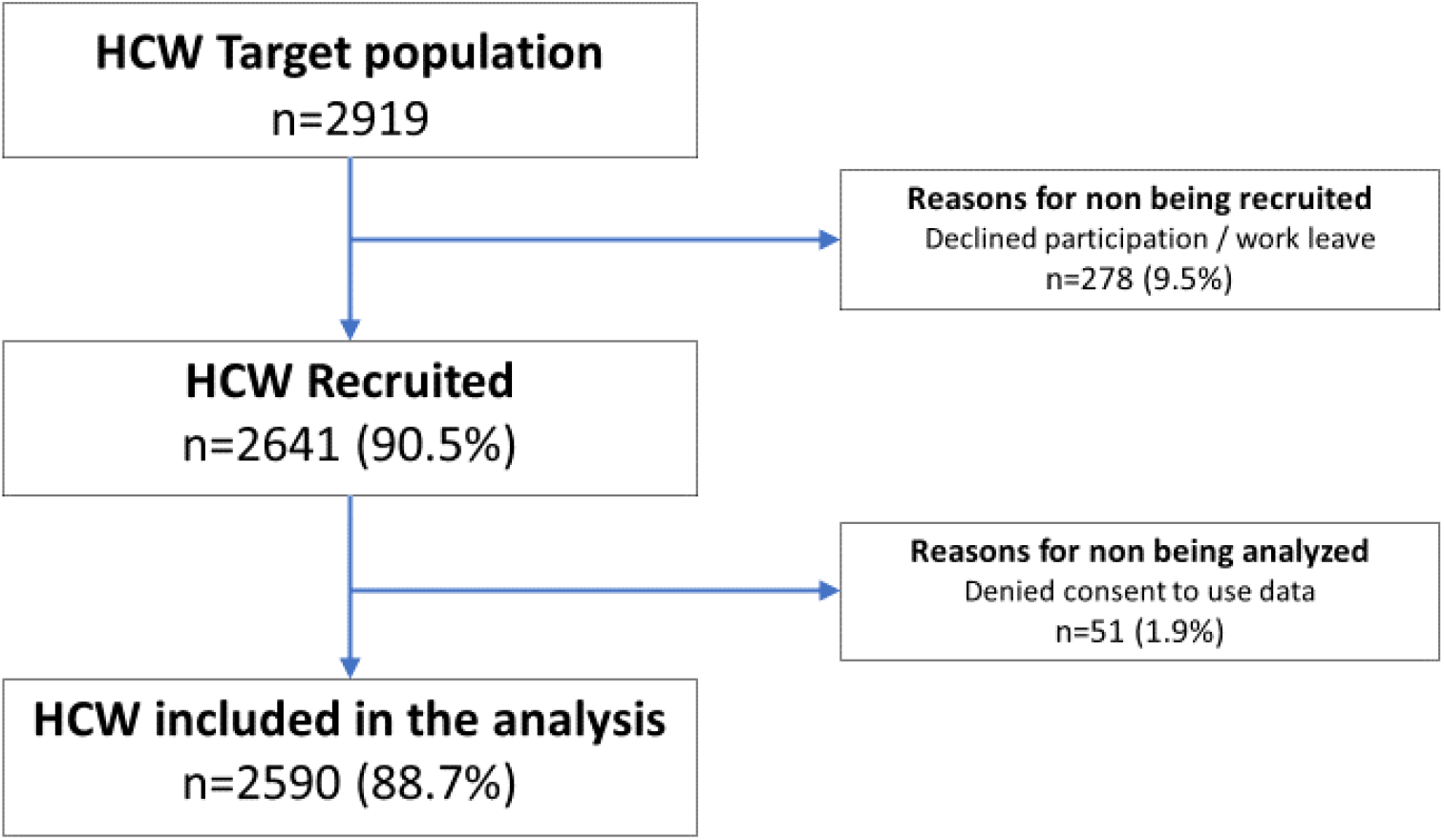
Flowchart of the study.

**Table 1.** Demographics. General description of the participants in the study. SD: standard deviation.

| VARIABLE |  | Total<br>n=2,590 (%) |
| --- | --- | --- |
| Women |  | 1,915 (73.9%) |
| Age (mean±SD) years |  | 43.8 SD 11.1 |
| Clinical conditions | Tobacco use | 545 (21%) |
|  | Chronic lung disease | 214 (8.3%) |
|  | High blood pressure | 180 (6.9%) |
|  | Obesity | 151 (5.8%) |
|  | Other cardiovascular disease | 68 (2.6%) |
|  | Diabetes mellitus | 54 (2.1%) |
|  | Immunodeficiency | 26 (1.0%) |
|  | Cancer | 9 (0.3%) |
|  | Liver disease | 7 (0.3%) |
|  | Chronic kidney disease | 8 (0.3%) |
|  | Pregnancy | 8 (0.3%) |
| Occupational SARS-CoV-2 exposure |  | 1,946 (75.1%) |
| Previous PCR test |  | 727 (28.1%) |
| Professional category | Technicians | 192 (7.4%) |
|  | Administrative and management | 170 (6.6%) |
|  | External workers | 337 (13%) |
|  | Patient carriers | 168 (6.5%) |
|  | Nurse | 687 (26.5%) |
|  | Physician | 564 (21.8%) |
|  | Nurse assistant | 472 (18.2%) |
| Work area | Critical care unit | 226 (8.7%) |
|  | External workers | 204 (7.9%) |
|  | Hospitalized COVID area | 887 (34.2%) |
|  | Hospitalized non COVID area | 373 (14.4%) |
|  | Management | 226 (8.7%) |
|  | Non-hospitalized non COVID area | 122 (4.7%) |
|  | Central units | 298 (11.5%) |
|  | Emergency Room | 253 (9.8%) |

A total of 2,369 (91.5%) participants reported some degree of direct exposure to SARS-CoV-2. The exposition was occupational in 1,946 (75%), contact with affected colleagues in 1,710 (66%), and non-occupational in 290 (12.2%) HCW. Among HCW with occupational exposition, 72% of them referred adequate PPE use.

### IgG results

Overall, SARS-CoV-2 IgG was positive in 818 HCW (31.6%), negative in 1,743 (67.3%) and borderline in 29 (1.1%). There were no differences IgG seropositivity positive for sex (31.6% women vs 33% men, p = 0.482) or age (positive 43.9 years [11.4 SD]) vs negative 43.6 years [11.2 SD]), p = 0.719) respectively.

### IgG results by area and professional category

High and medium risk areas had higher rate of seropositivity (33.1%, [450/1,359] and 33.8% [257/760]) than low risk areas (25.8%, [48/201]), p = 0,007 (figure 2). The proportion of seropositive HCW among working areas was: ER (32.8%, 83/253), critical care (23.8%, 53/223), COVID-19 admitted patients (35.5%, 311/875), non-COVID-19 admitted patients (38.3%, 141/368), non-COVID-19 clinical care units (32,8%, 40/122) central units (29%, 85/293) administrative and management areas(24.9%, 56/225) and external workers 23.9%, 48/201; p<0.001 (table 2)

**Table 2.**
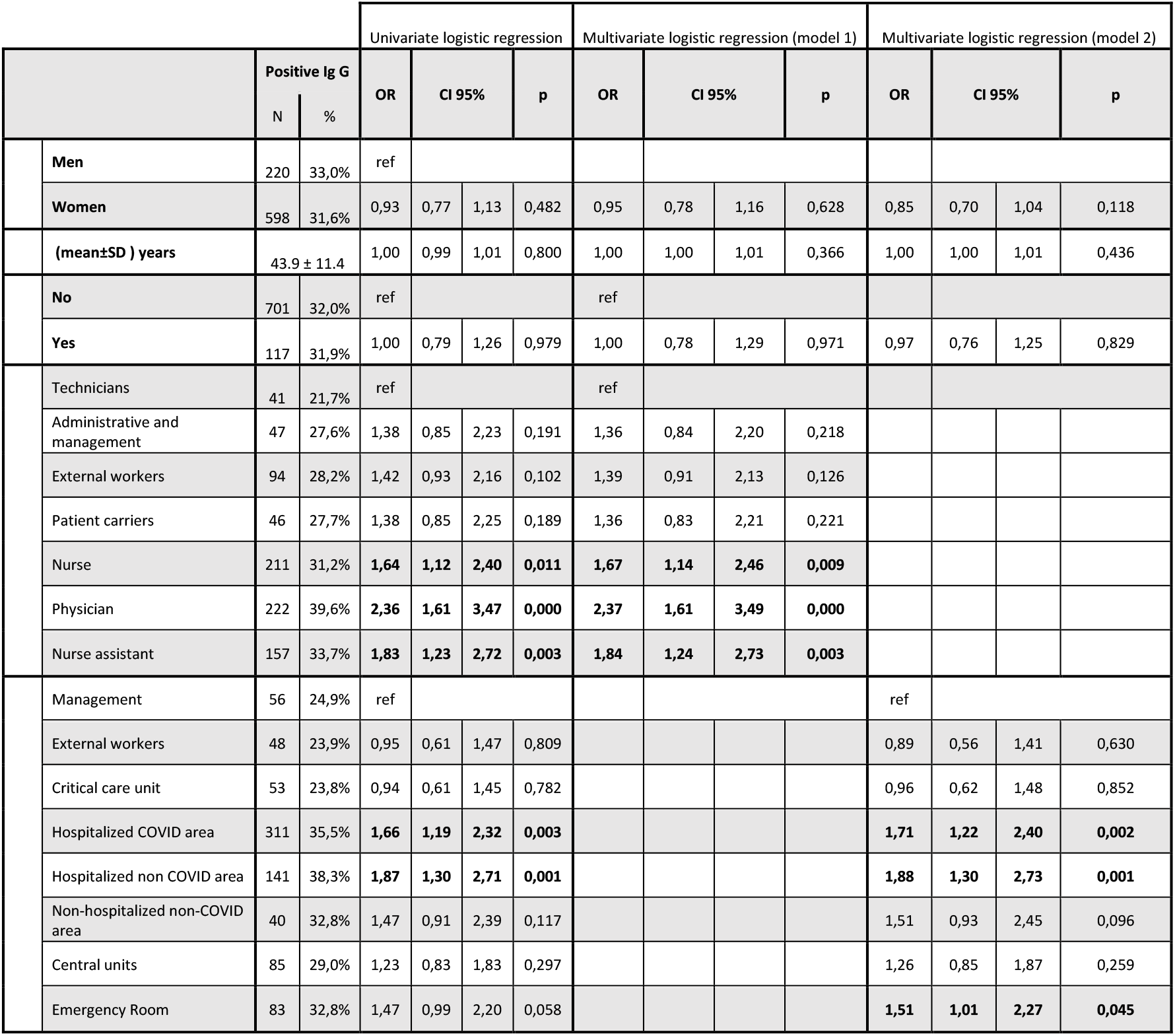
Univariate and multivariate analysis. First model included sex, age, cardiovascular disease and professional category and the second model included sex, age, cardiovascular disease and work area. OR: odds ratio; CI95% confidence interval 95%. Central units included pharmacy, laboratory, radiology, pathology. External workers included cooks, food service, cleaners, ambulance drivers, store sellers, and watchmen.

**Figure 2.**
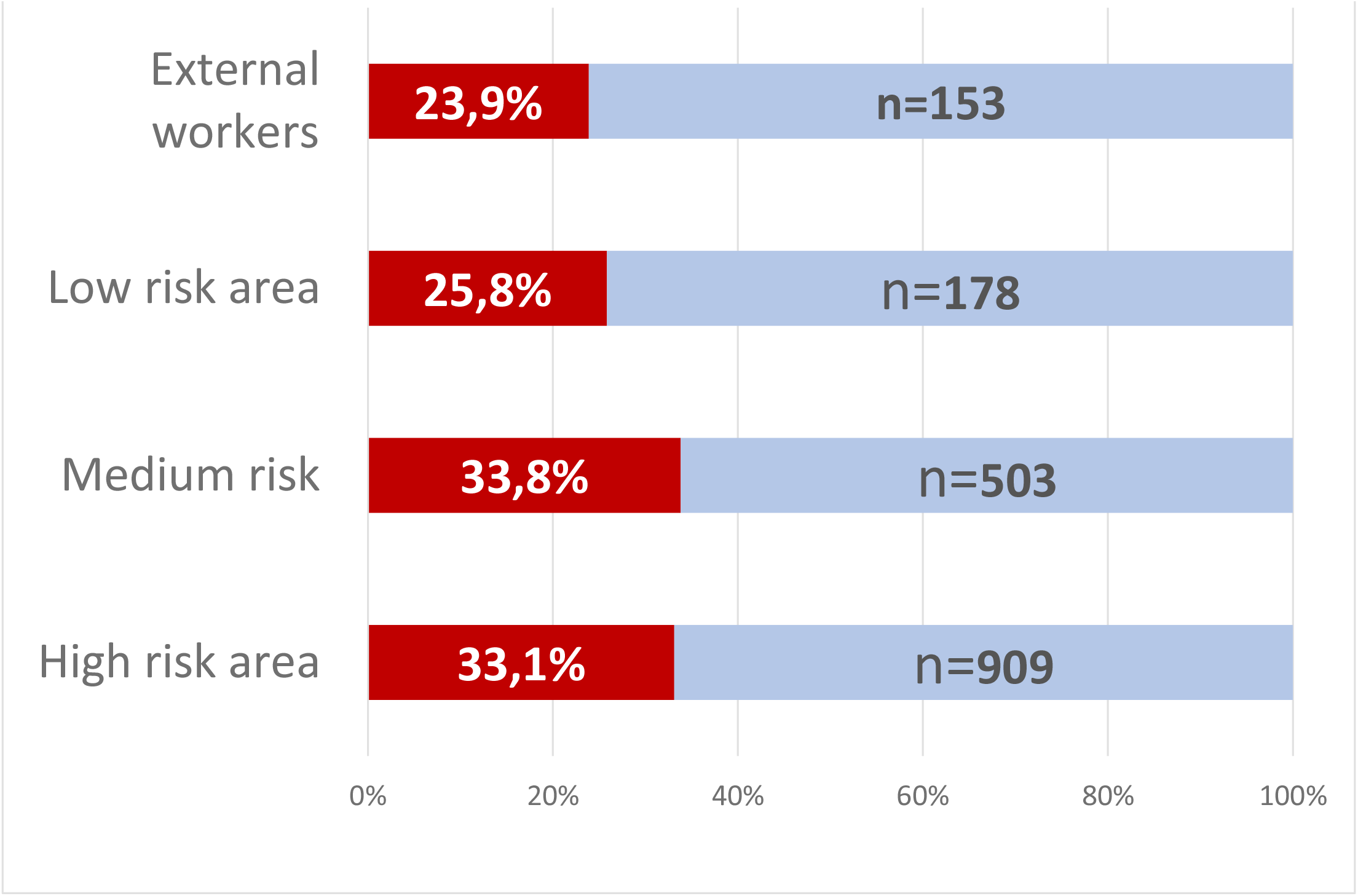
Distribution of SARS-CoV-2 seropositivity by risk area. Red bars: percentage positive IgG HCW. Blue bars: percentage of IgG negative HCW. Numbers of negative HCW are shown over the blue bars. High risk areas of exposure to SARS-CoV2: included professionals with direct contact with COVID-19 patients: critical care & anesthesiology, ER and COVID- hospitalization ward. Medium risk areas included professionals attending patients not suspected having COVID-19 both in hospital wards and outpatient clinics, and central units (pharmacy, laboratory, radiology, pathology). Low risk was attributed to administrative and management units. External workers included cooks, food service, cleaners, ambulance drivers, store sellers, and watchmen. Chi square, p = 0,007

Physicians were the most infected professional category, (39.6%, 222/561) followed by nurse assistant, (33.7%, 157/466), nurses (31.2%, 211/676), external workers (28.2%, 94/333), patient carrier (27.7%, 46/166), administrative and management staff (27.6%,47), and finally technicians (21.7%, 41/170), p< 0.001 (table 2).

Participants who referred use of inappropriate PPE were 27.0%. The rate of seropositivity among them was 42.0% (219/522) as compared to 27.7% (374/1354) in cases with referred appropriate PPE use, p< 0.001. However, this difference disappeared when the sample was stratified by previous attention to the OHU.

### Symptoms

A total of 397 out of 818 (48,5%) IgG positive HCW did not consult at the OHU previous to the seroprevalence study, so they did not consider that their symptoms, if any, could be related to SARS-CoV-2 infection. However, among them, 193 (48.6%) recalled minor symptoms in the study interview that they had not attributed to potential SARS-CoV-2 infection.

On the contrary, during the period of COVID-19 clinical care, a total of 421 out of 818 HCW (51.4%) attended to the OHU because of symptoms suggestive of COVID-19 and were tested by PCR for SARS-CoV-2; among them, 306/421 (72.7%) tested positive. A small proportion ((48 cases/421 (11,4%%)) of HCW were further evaluated at the ER, and 25 (5,9% [25/421]) required hospital admission (figure 3). PCR tests were not performed in asymptomatic individuals.

**Figure 3.**
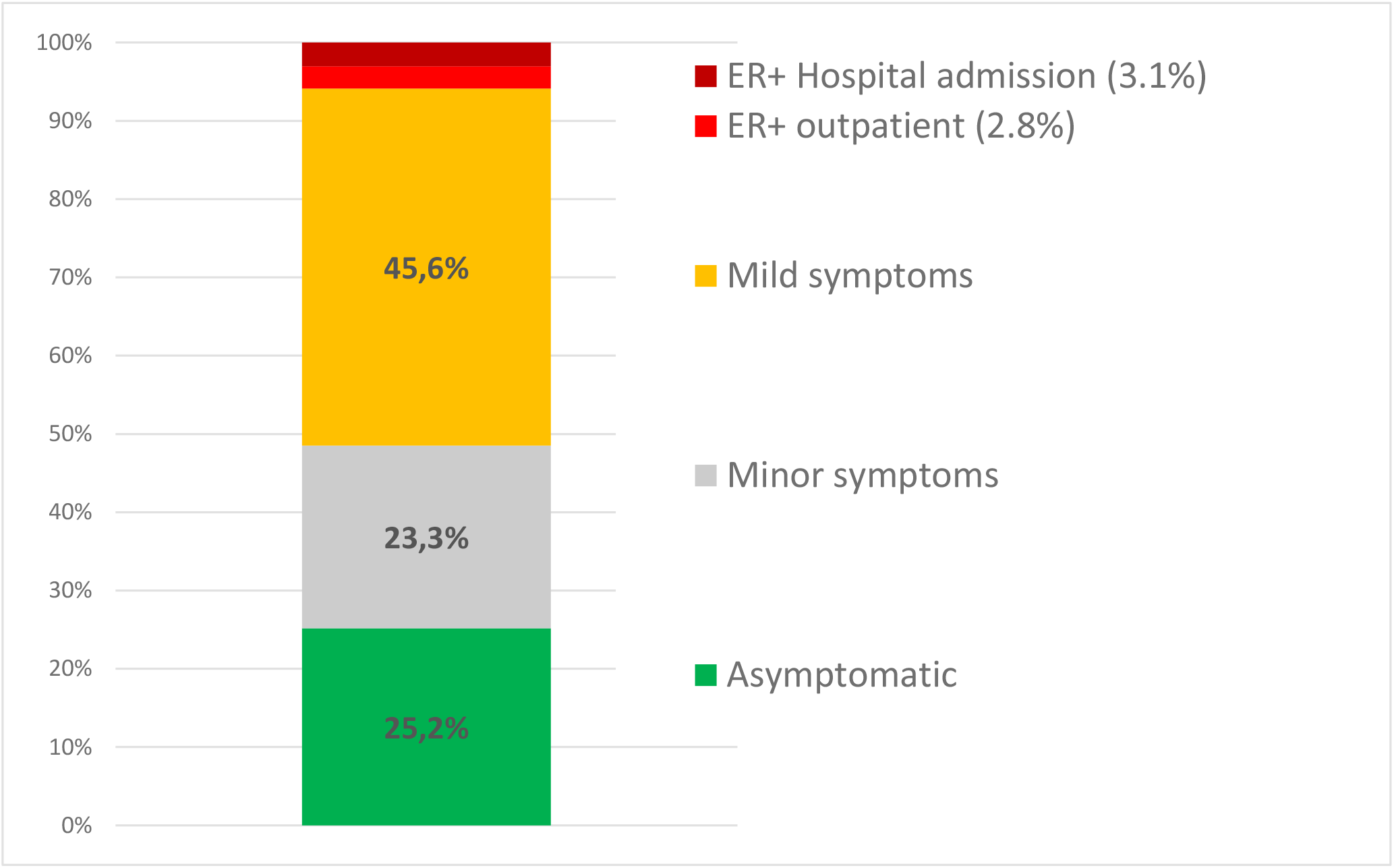
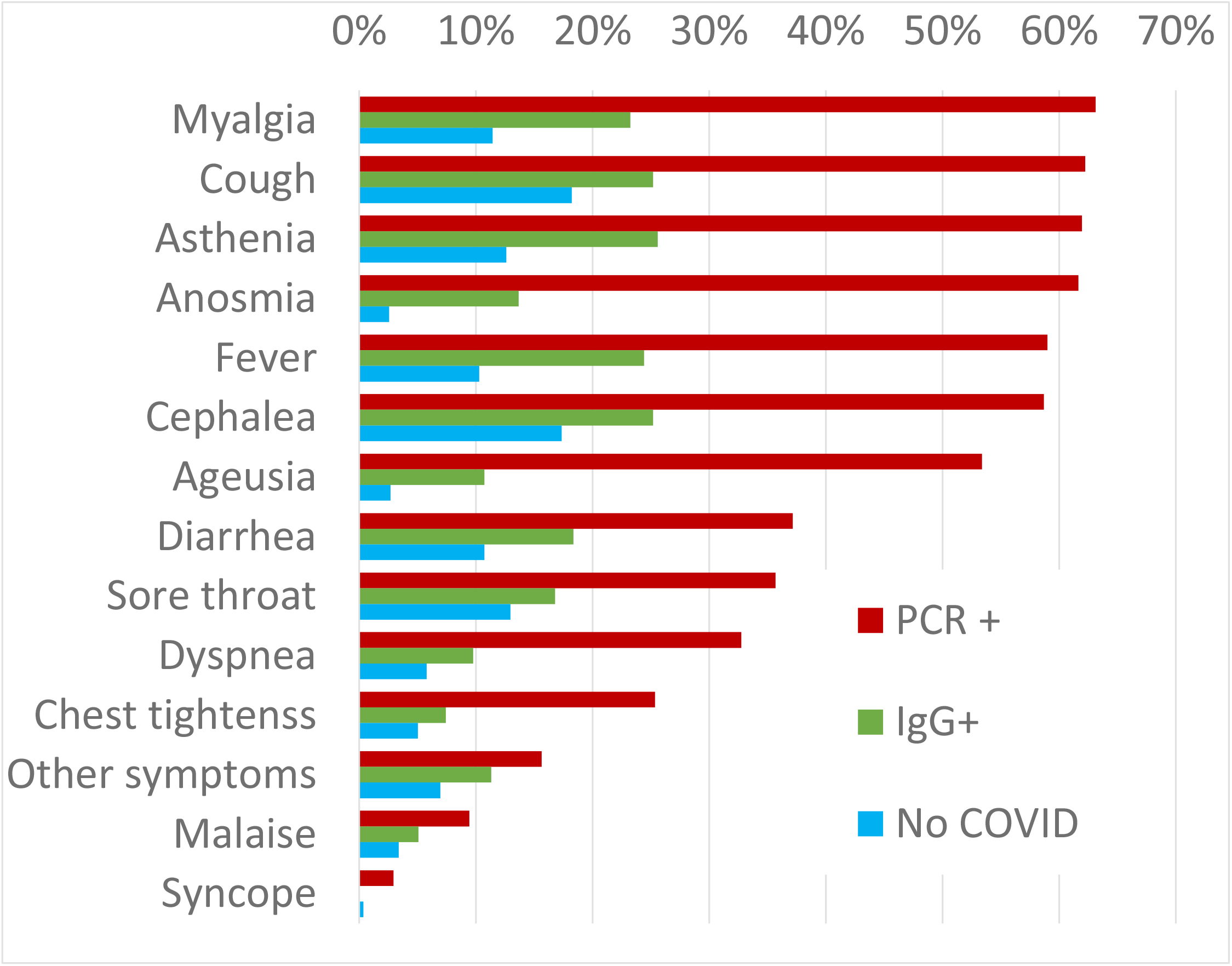
Clinical symptoms. Panel A. Clinical spectrum of COVID-19 in the hospital. ER: Emergency Room. Panel B. Symptoms description of HCW by diagnosis. PCR: HCW diagnosed by positive PCR. IgG: HCW diagnosed just by serology. No COVID: HCW with negatives PCR and IgG.

The most common symptoms among PCR-confirmed COVID-19 cases were myalgia (63.1%), cough (62.2%), asthenia (61.9%), anosmia (61.7%), fever (59.0%), cephalea (58.7) and ageusia (53.4%) (figure 3). Those HCW with COVID-19 diagnosed Symptoms any, could be related to SARS-CoV-2 infection. However, among them, 193 (48.6%) solely by serology presented symptoms less frequently. Of note, some symptoms were also reported among HCW without COVID-19.

Forty-two out of 818 IgG positive HCW referred mild symptoms in the 14 days previous to study evaluation and were tested by PCR for SARS-CoV2. Among them 8 HCW were positive and they were sent home for quarantine.

### PCR results

Three hundred and six, 90.8% (306/339) of HCW with previous positive PCR were SARS-CoV-2 IgG positive. Conversely, 30.2% (115/388) of HCW with negative SARS-CoV-2 PCR had positive IgG. Finally, 9.2% (31/339) with positive PCR had a negative IgG result. Median time from positive PCR and negative IgG test was 21 days (IQR 14–26).

We evaluated factors associated with SARS-CoV-2 seropositivity by logistic regression analysis. The independent variables included in the model were age, sex, cardiovascular disease, professional category (model 1) and work area (model 2). HCW with a significant increased probability of SARS-CoV-2 IgG positive were physicians (OR 2.37, CI95% 1.61–3.49) nurse (OR 1.67, CI95% 1.14–2.46), and nurse assistant (OR 1.84, CI95% 1.24–2.73), and HCW that works at COVID-19 hospitalization areas (OR 1.71, CI95% 1.22–2.40), non-COVID-19 hospitalization areas (OR 1.88, CI95% 1.30–2.73), and at ER (OR 1.51, CI95% 1.01–2.27) (table 2)

## DISCUSSION

This is the first study of the SARS-CoV-2 seroprevalence of all HCW, regardless whether they were or not direct employee of the hospital. We found a relatively high proportion (30%) of HCW with a positive IgG for SARS-CoV-2. A recent study from Spanish population showed a national prevalence of 5%, but 11% in Madrid^23^. A partial explanation for a higher prevalence at our hospital is the higher exposition to the virus in the city of Alcorcón. Data from Madrid Regional Government shows that Alcorcón had a slightly higher incidence of COVID-19 than the region of Madrid^24^. Furthermore, a recent study from a large hospital in Barcelona showed a prevalence in a sample of HCW of 11.6%, doubling the seroprevalence of the general population^25^, strengthening the notion of hospitals as a places of risk for SARS-Co2 infection among workers. Unsurprisingly, external workers (23.9%) and non-clinical workers (25.8%) had lower seroprevalence than average^26^, although still much higher than the general population in Madrid^23^. These data suggest a role for nosocomial transmission also for non-clinical workers^27,28^

Regarding clinical workers (all of them direct employees of the hospital), the rate of positive IgG was virtually identical among workers with direct contact with COVID-19-patients and those taking care of non-COVID-19 patients, as it has been reported in other settings^20^. Some have proposed that workers with no direct contact with COVID-19 could have been infected in the population (in a context in which the actual seroprevalence in the population was unknown)^20^. Our data argue against it: clinical worker in non-COVID areas become seropositive likely because of in-hospital contact, either from asymptomatic patients or colleagues.

These data suggest that the non-COVID-19 clinical areas are indeed an unrecognized potential source for COVID-19 infection among workers^27^. A recent meta- analysis estimates that nosocomial transmission is the source of SARS-CoV-2 infection in about 44% of cases^29^. This estimation is further increased up to almost 90% of cases in a mathematical model.^30^

Since universal COVID-19 screening has not been a usual practice implemented it is conceivable that a substantial proportion of so-called non-COVID-19 patients may be actually subclinical or unnoticed COVID-19 cases^31,32^. Our results are in agreement with a high rate of nosocomial transmission reported among workers in a dialysis unit in New York^33^. These data emphasize the need for universal screening of all in-hospital patients as recommend World Health Organisation^34–36^ and we are already implementing.

A similar proportion of seropositivity among clinicians taking direct care of COVID-19 patients suggest that the isolation protocols and PPE appear sufficient to prevent high levels of nosocomial transmission in our setting^13,37^. Of note, critical care workers had one of the lowest seropositivity rates in our study. Indeed, our hospital prioritized the use of the best available PPE for critical care units, where virtually all patients were COVID-19 at the peak of the epidemic^38^.

The clinical spectrum of COVID-19 in our workers resembles that described for the general population: about half of them are asymptomatic or paucisymptomatic^39–43^ and less than 60% had fever (figure 3). That means that most infected workers remain undetected unless there is a universal screening^44,45^. In retrospect, about 50% of seropositive workers attending to the serology study recalled minor symptoms that did not prompt a request for OHU evaluation. Thus, only about one fourth of IgG positive workers were fully asymptomatic, as reported in other studies^31,33^.

Regarding workers with overt symptoms suggesting COVID-19 disease most of them (83%) had a mild disease that could be managed in the outpatient setting. About 6% required ER visit and 3% required hospital admission. There were no deaths. This is hardly surprisingly since there is no elder population among active workers^10^.

To prevent nosocomial transmission both patients and health care workers should be screened for SARS-CoV-2 infection regardless of the absence of typical symptoms for COVID-19 disease^45^ as asymptomatic transmission is being increasing recognized as very relevant in SARS-CoV-2 spread.^9,27,46^.

Our study has some limitations that deserve consideration. First, we do not have data about Ig M or concurrent PCR. However, our study was designed to have a picture of past exposure to the virus in all our workers. We did not pursue an evolutionary perspective of the disease. Second, the samples were collected over two weeks, so the interpretation of the prevalence must be related to the average prevalence at that time.

Nonetheless, our work has several strengths. First, the quality of the technology we had used seems to be one of the highest sensitivities available (ELISA)^47–49^. Second, we had a virtually universal representation of all workers of the hospital (90%), including external employees, an evaluation hardly performed. Additionally, we identified the particular function of all employees in a time of changing roles for clinicians in the middle of the crisis. In addition, its close temporal vicinity with the serologic study in the Spanish population allows for a direct comparison.

In conclusion, seroprevalence unmasked a high rate of infection previously unnoticed in HCW. Clinical care of COVID-19 unscreened patients is associated with a similar prevalence of SARS-CoV-2 antibodies as the one found in COVID-19 facilities uncovering a relevant source for nosocomial SARS-CoV-2 transmission. In addition, apparently healthy HCW may also be another relevant source for SARS-CoV-2 transmission. HCW testing could reduce in-hospital transmission^50^. Serosurveys in hospitals may be helpful to design strategies to control SARS-CoV-2 epidemic.

## Data Availability

Individual data will be available upon request

## Acknowledgment

We thank all workers of the Hospital Universitario Fundación Alcorcón, who bravely and generously faced the COVID-19 epidemic during the months of March and April for their everyday work and cooperation in this study.

## SUPPLEMENT 1

Previous health conditions: tobacco use, hypertension, obesity, cardiovascular disease, chronic liver disease, chronic lung disease or asthma, chronic renal failure, immunodeficiency, or pregnancy.

COVID-19 related symptoms: fever, myalgia, cough, sputum, dyspnea, rhinorrhea, sore throat, diarrhea, anosmia/hyposmia, ageusia/dysgeusia, asthenia, chest pain, headache, syncope, others), SARS-CoV-2 PCR test result as well as the severity of disease when appropriate (out-patient evaluation, ER consultation, hospital admission and clinical outcome.

## SUPPLEMENT 2

We measured serum IgG antibody by an enzyme-linked immunosorbent assay (ELISA) IgG2 using a SARS-CoV-2 S spike and Nucleocapsid recombinant antigens (Diapro (Palex), Italy), to screen for the presence of human anti- SARS-CoV-2 IgG. This assay (CE approved) was used according to the manufacturer’s protocol. Reported sensitivity of the assay by the manufacturer was 98%.

The results of the tested samples were determined by calculating the ratio of the optical density (OD) value of the sample to the OD value of the cut-off. (Co) Ratios ≥ 1.1 were considered positive, ratios ≥ 0.9 to < 1.1 were considered borderline, and ratios < 0.9 were considered negative. All assays were run following manufacter’s instructions on the platforms DSX System (Palex Medical SA) and Triturus (Grifols Movaco SA).

Sensitivity of the assay using samples from 337 workers from our series with results previous positive PCR was 90.8% (manufacture shows 98%). Specificity manufacture’s instructions shows that the assay was tested on hundreds of samples collected before the outbreak of COVID-19. A value of > 90% was found.

Index values considered “borderline” were tested on Strips-module Enzyme Immunoassay for the confirmation of IgG antibodies to COVID-19–19 major antigens. This assay detects IgG antibodies against the SARS-CoV-2: Spike glycoprotein 1, Spike glycoprotein 2 and nucleocapside proteins. A sample is considered for a certain antibody negative S/Co< 1, equivocal 1 < S/Co< 1.2, positive S/Co>1.2. These samples were run on the platform DSX System (Palex Medical SA). The manufacter’s instructions shows that the assay was tested on hundreds of samples collected before the outbreak of COVID-19–19. A value of > 98% was found. About 2% of the reactive “normal” population shows a reactivity to Nucleocapsid. A first minimum study carried out in the context of a Public Health Emergency on samples from a cohort of infected patients showed a sensitivity of about 98%.

This assay detects IgG antibodies against the SARS-CoV-2 independently : Spike glycoprotein 1, Spike glycoprotein 2 and nucleocapside proteins. A sample is considered positive S/Co>1.2. Specificity was found > 98% and sensitivity of about 98%. The internal validation was performed by correlation with previously evaluated PCR test as gold standard.

For molecular diagnosis of SARS-CoV-2 infection of nasopharyngeal swabs were processed by automatized extraction using the MagNa Pure Lc instrument (Roche Applied Science, Mannheim, Germany) and real time reverse transcription polymerase chain reaction using the SARS-Cov-2 nucleic acid detection Viasure kit (CerTest Biotec S.L.), following the manufacturer’s instructions.

For this rRT-PCR, we used Bio-Rad CFX96™ Real-Time PCR Detection System. We amplified two different viral regions: ORF1ab gene is amplified and detected in FAM channel, and N gene is amplified and detected in ROX channel and the internal control (IC) in HEX cannel.

Cycle threshold values, i.e., number of cycles required for the fluorescent signal to cross the threshold in rRT-PCR, were quantified viral load, with lower values indicating higher viral load. A sample was considered positive when the rRT-PCR Ct value was ≤40. Positive and negative control were included in each run for each assay.

